# A Collaborative Approach to Improving Missense Mutational Effect Predictions in Oncoproteins

**DOI:** 10.1101/2025.05.15.25327738

**Authors:** Ayten Dizkirici Tekpinar, Mustafa Tekpinar

**Affiliations:** Department of Molecular Biology, Faculty of Science, Van Yüzüncü Yıl University, 65080, Van, Türkiye; Department of Physics, Faculty of Science, Van Yüzüncü Yıl University, 65080, Van, Türkiye

**Keywords:** Missense mutations, oncoproteins, variant effect prediction, cancer genomics

## Abstract

Missense mutations in oncoproteins play a key role in cancer, which makes their accurate classification essential for cancer diagnosis and treatment. Although various variant effect prediction algorithms have been developed, their accuracy varies across different proteins. In this study, we introduce the APE score, a novel ensemble metric that integrates three state-of-the-art unsupervised predictors—AlphaMissense, PRESCOTT, and ESM1b. We evaluated APE score’s performance using nine deep mutational scanning (DMS) experiments on five oncoproteins and a dataset of 1068 clinically labeled variants from 24 oncoproteins. Our results show that APE score is better than its individual components in six out of nine DMS experiments and achieves superior accuracy in distinguishing pathogenic variants from benign ones. Additionally, we established classification thresholds to categorize 444087 variants into ‘likely pathogenic,’ ‘variants of unknown significance (VUS),’ and ‘likely benign’ categories. The APE score reduces misclassification rates and provides a more reliable tool for assessing the functional impact of missense mutations in cancer-related proteins. All scores and classifications are provided in a publicly available repository to support further research and clinical applications.

## Introduction

Cancer is a disease affecting millions of people worldwide (Bray et al. 2024). All cancers are caused by genetic changes, which can be inherited or acquired over a lifetime (Pearson and Van der Luijt 1998). As a result, correct identification of impact of genetic changes such as missense mutations in oncoproteins is of utmost importance both for cancer diagnosis and treatment. Despite extensive clinical and experimental efforts to identify impact of missense mutations in oncoproteins, only a limited number of variants in oncoproteins have been identified. Due to this reason, fast and accurate variant effect prediction algorithms are highly needed in oncology as well as in human genetics generally.

We have witnessed a significant progress in variant effect prediction algorithms in recent years (Livesey and Marsh 2023; Livesey and Marsh 2024). Increasing amount of genetic data is an important contributor to this progress. Another major contributor is machine learning algorithms. In particular, deep learning methods started to change the variant effect prediction landscape in addition to protein structure prediction field. AlphaMissense (Cheng et al. 2023) is one of the recent state-of-the-art variant effect prediction algorithms that utilizes deep learning and revolutionary protein structure prediction algorithm AlphaFold (Jumper et al. 2021). Even though it is not a deep learning algorithm, PRESCOTT can reach accuracy of AlphaMissense or exceed its performance for many proteins (Tekpinar et al. 2025). ESM1b is another recent variant effect prediction algorithm that uses protein language models to predict missense mutational effects (Brandes et al. 2023). Despite a significant progress in prediction performance of variant effect prediction algorithms, almost all of the variant effect prediction algorithms, including the mentioned ones, fail to predict missense mutation effects for numerous clinically/experimentally verified variants. Accuracy level can vary significantly from protein to protein and average Spearman correlation is about 0.500 currently (see a detailed comparison of many methods at https://proteingym.org/benchmarks) (Notin et al. 2023). Hence, it is evident that there is a large room for improving variant effect prediction algorithms.

Generally, every mutational effect prediction method tries to establish its superiority to the available methods, sometimes from different perspectives. In this work, we adopt a collaborative approach instead of the common competitive approach. We build a new score called APE score from three unsupervised state-of-the-art methods, namely **A**lphaMissense, **P**RESCOTT and **E**SM1b. We investigate APE score accuracy with Spearman correlation on nine deep mutational scanning experiments conducted on 5 oncoproteins. Then, we test APE score performance for distinguishing pathogenic variants from benign variants on a set of 1068 pathogenic or benign labeled variants in 24 oncoproteins. After establishing that APE score is better than its individual components, we use it for classification of all variants in 24 oncoproteins into ‘likely pathogenic’, ‘variants of unknown significance (VUS)’ and ‘likely benign’ classes. Finally, we release APE score and classifications for 444087 variants in 24 oncoproteins in a public repository to the service of scientific and clinical community.

## Materials and Methods

### A Set of 24 Cancer Proteins

We used a previously used dataset of 24 oncoproteins that we will call ‘large dataset’ throughout the paper (Keskin Karakoyun et al. 2023). The large dataset contains 1068 binary labeled (pathogenic or benign) variants, of which 438 are pathogenic mutations and 630 are benign mutations (Fig. 1 A). A small set of 19 proteins containing 267 pathogenic and 373 benign variants was selected from the large dataset for further analysis. The small dataset serves two purposes: 1) It helps us to check validity of our claims if some large proteins are pruned from the dataset. 2) It gives us a chance to include additional algorithms because all algorithms presented here do not have results for all of the proteins in the large dataset.

**Fig. 1.**
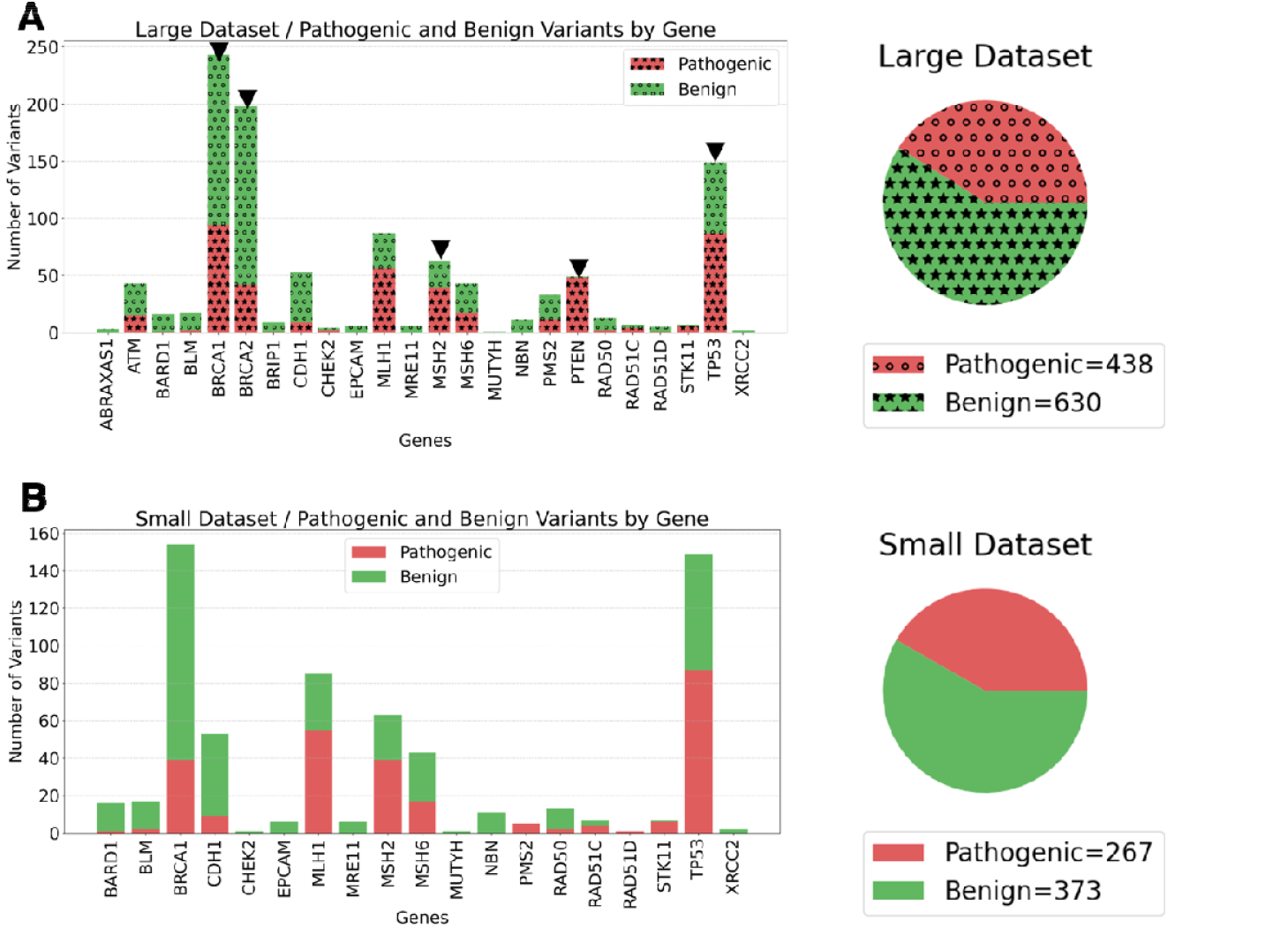
The large and small datasets containing oncoproteins investigated in this study. A) Names of genes and number of variants for the large dataset (24 proteins) are given as a bar plot in the left panel. Distribution of pathogenic (red area with circles) and benign (green area with filled stars) variants for the large dataset is given as a pie chart in the right panel. Black downward triangles highlight proteins with at least one deep mutational scanning experiment. B) Names of genes and number of variants for the small dataset (19 proteins) are given as a bar plot in the left panel. Distribution of pathogenic (red area) and benign (green area) variants for the small dataset is given as a pie chart in the right panel.

### Obtaining Predictions of Different Methods

AlphaMissense (Cheng et al. 2023) results were downloaded from https://console.cloud.google.com/storage/browser/_details/dm_alphamissense/AlphaMissense_aa_substitutions.tsv.gz. ESCOTT (Tekpinar et al. 2025) results were download from ESCOTT database at http://prescott.lcqb.upmc.fr/prescott_data_main.php. Allele frequencies of the proteins were downloaded from gnomAD (https://gnomad.broadinstitute.org/) and PRESCOTT scores were calculated using version 4.0.0 of gnomAD data (Karczewski et al. 2020). ESM1b (Brandes et al. 2023) data was downloaded from https://huggingface.co/spaces/ntranoslab/esm_variants. GEMME (Laine et al. 2019) results for our proteins were extracted from https://datadryad.org/dataset/doi:10.5061/dryad.vdncjsz1s (Abakarova et al. 2023). PolyPhen2 (Adzhubei et al. 2010) web server at http://genetics.bwh.harvard.edu/pph2/bgi.shtml was used to calculate PolyPhen HumVar and HumDiv scores. Sift (Ng and Henikoff 2003) server at https://sift.bii.a-star.edu.sg/www/SIFT_seq_submit2.html was used for Sift score computations. VARITY (Wu et al. 2021) scores were downloaded from http://varity.varianteffect.org/. gMVP (Zhang et al. 2022) data was obtained from a precalculated dataset deposited at https://www.dropbox.com/scl/fi/crjpntg530gucq8ch66y2/gMVP.2021-02-28.csv.gz?rlkey=kmg7qsomnunjitgrzej5gexmh&e=1&dl=0. Finally, popEVE (Orenbuch et al. 2023) results for each protein were downloaded from https://pop.evemodel.org/.

### Deep Mutational Scanning (DMS) Experiments

Unfortunately, there are DMS experiments for only 5 proteins out of 24 proteins in the large dataset. Nine experiments have been conducted on these 5 oncoproteins and they were downloaded from ProteinGym v1.0.0 website at https://proteingym.org/download (Notin et al. 2023).

### APE Score Calculation

APE score is a linear combination of AlphaMissense, PRESCOTT and ESM1b. AlphaMissense and PRESCOTT scores are already in [0.0, 1.0] range where 0.0 indicates low mutational impact and 1.0 indicates high mutational impact, namely a potentially pathogenic impact. ESM1b scores generally vary between [-20, 0] and low scores indicate high impact. ESM1b raw scores need to be rescaled between zero and one for compatibility with AlphaMissense and PRESCOTT. Minmax normalization was applied to each protein after calculation of all variants because minmax normalization doesn’t change the distribution of the scores. All protein context was taken into account during minmax normalization. Then, all minmax normalized ESM1b scores were subtracted from 1 to ensure scores close to one denote high mutational impact. Eventually, APE scores were calculated as follows:

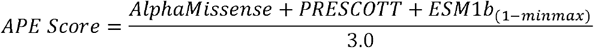

APE score is a quantity between zero and one. The scores close to zero indicate low mutational impact while high impact mutations have APE scores close to one.

## Results and Discussions

### Prediction Performance of APE Score for Deep Mutational Scanning Experiments

There are several deep mutational scanning experiments for five of the proteins in our dataset (see proteins with downward black triangles in **Fig. *1*** A). We measured Spearman correlation of APE scores and three state-of-the-art methods (AlphaMissense, PRESCOTT and ESM1b) with deep mutational scanning experiments to quantify prediction performance. Spearman correlation of APE score with BRCA1_HUMAN_Findlay_2018 experiment (Findlay et al. 2018) is 0.599 and it exceeds individual components of APE score (**Fig. *2*** A). Interestingly, ESM1b becomes the best predictor for BRCA2_HUMAN_Erwood_2022_HEK293T (Erwood et al. 2022) experiment while APE score follows it with a Spearman correlation of 0.455 (**Fig. *2*** B). Even though APE score is not the best for BRCA2, it is still better than AlphaMissense and PRESCOTT performance. It is interesting to note that this experiment contains only 265 variants, which is the experiment with the smallest number of variants, despite gigantic size (3418 amino acids) of the protein (see Table S1 for details). MSH2 is another important protein involved in cancer with DMS data in ProteinGym. APE scores have 0.426 Spearman correlation with MSH2_HUMAN_Jia_2020 experimental data (Jia et al. 2021) and it surpasses the other three constituent methods (**Fig. *2*** C). Both of PTEN DMS experiments (Mighell et al. 2020; Matreyek et al. 2021) demonstrate that APE score outperforms its individual components in terms of Spearman correlation metric for this protein (**Fig. *2*** D). It is possible to analyze Spearman correlation performances of pairwise combinations of AlphaMissense, PRESCOTT and ESM1b instead of triple combination as in APE score (Fig. S1). Double combinations become slightly better than the APE score in MSH2_HUMAN_Jia_2020 and PTEN_HUMAN_Mighell_2018 experiments (Mighell et al. 2020), while APE score is better in the others. Furthermore, results for four DMS experiments conducted on P53 tumor suppressor protein are presented in Fig. S2 and S3 of the Supplementary Information. APE score predictions for Kotler_2018 (Kotler et al. 2018) and Giacomelli_2018_Null_Etoposide (Giacomelli et al. 2018) experiments are better than the other methods, while APE scores have almost the same level of accuracy for the remaining two experiments. Pairwise combinations of AlphaMissense, PRESCOTT and ESM1b do not have a significant superiority compared to APE score in all P53 DMS experiments. To summarize, we demonstrated here that APE score outperforms its components in six out of nine DMS experiments and it has almost same level of accuracy in two experiments.

**Fig. 2.**
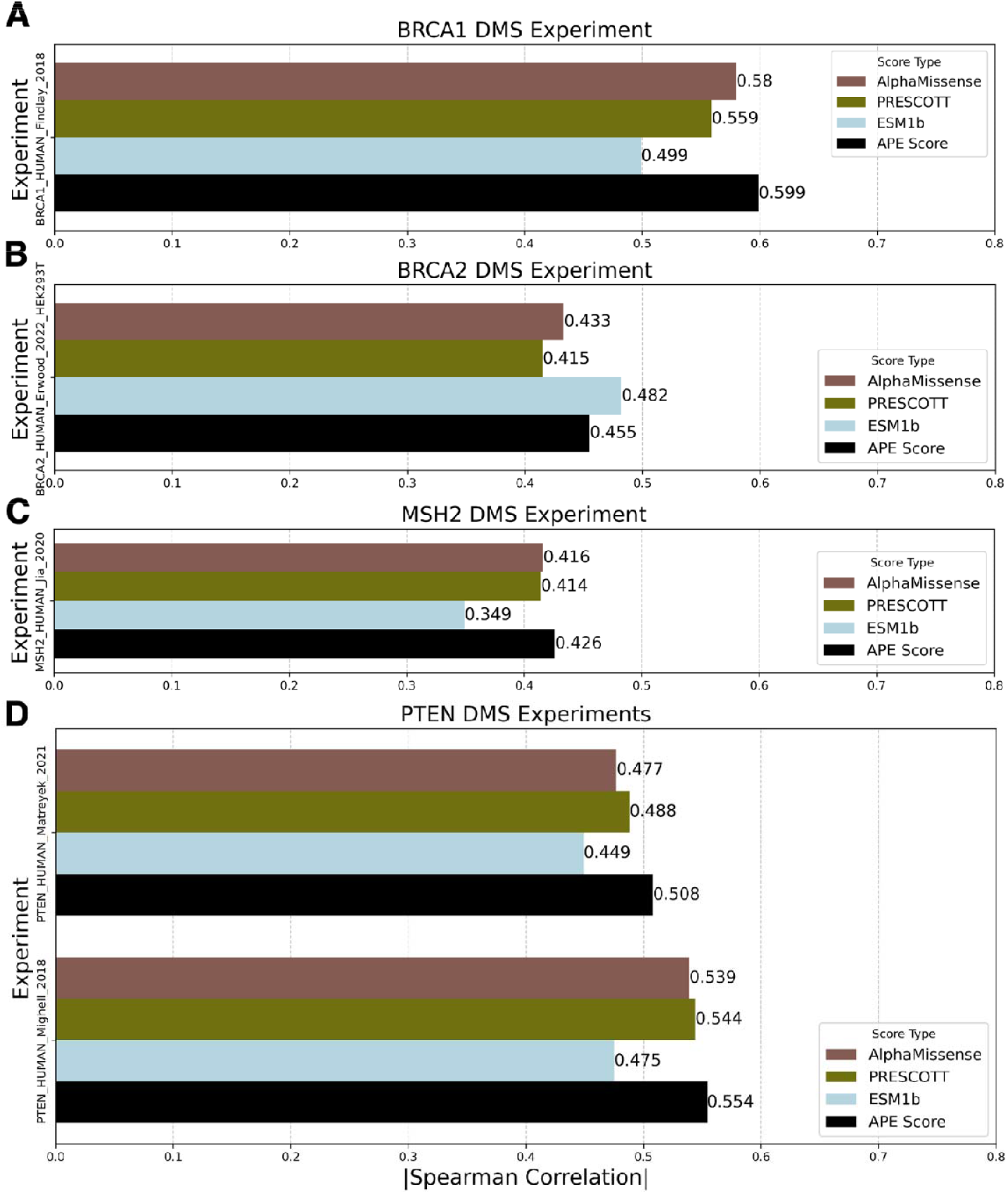
Comparison of AlphaMissense (brown bars), PRESCOTT (olive bars) and ESM1b (light blue bars) Spearman correlation performances with APE score performance across various deep mutational scanning experiments for A) BRCA1 B) BRCA2 C) MSH2 D) PTEN.

### Prediction Performance of Various Algorithms for Binary Labeled Mutants

We calculated prediction quality of nine methods for the variants in the large dataset with AUC and AUC-Precision-Recall (AUC-PR) metrics (Fig. 3 A), while prediction quality was investigated for sixteen methods in the small dataset (Fig. 3 B). If AUC values are considered, PRESCOTT becomes the best predictor with an AUC value of 0.958 in the large dataset. When assessed with AUC-PR, AlphaMissense is the best predictor in the large dataset. Top performer becomes a supervised prediction algorithm called VARITY in terms of both AUC and AUC-PR metrics in the small dataset. When VARITY derivatives are excluded, PRESCOTT (AUC=0.952) and AlphaMissense (AUC-PR=0.941) are the second-best predictors in the small dataset.

**Fig. 3.**
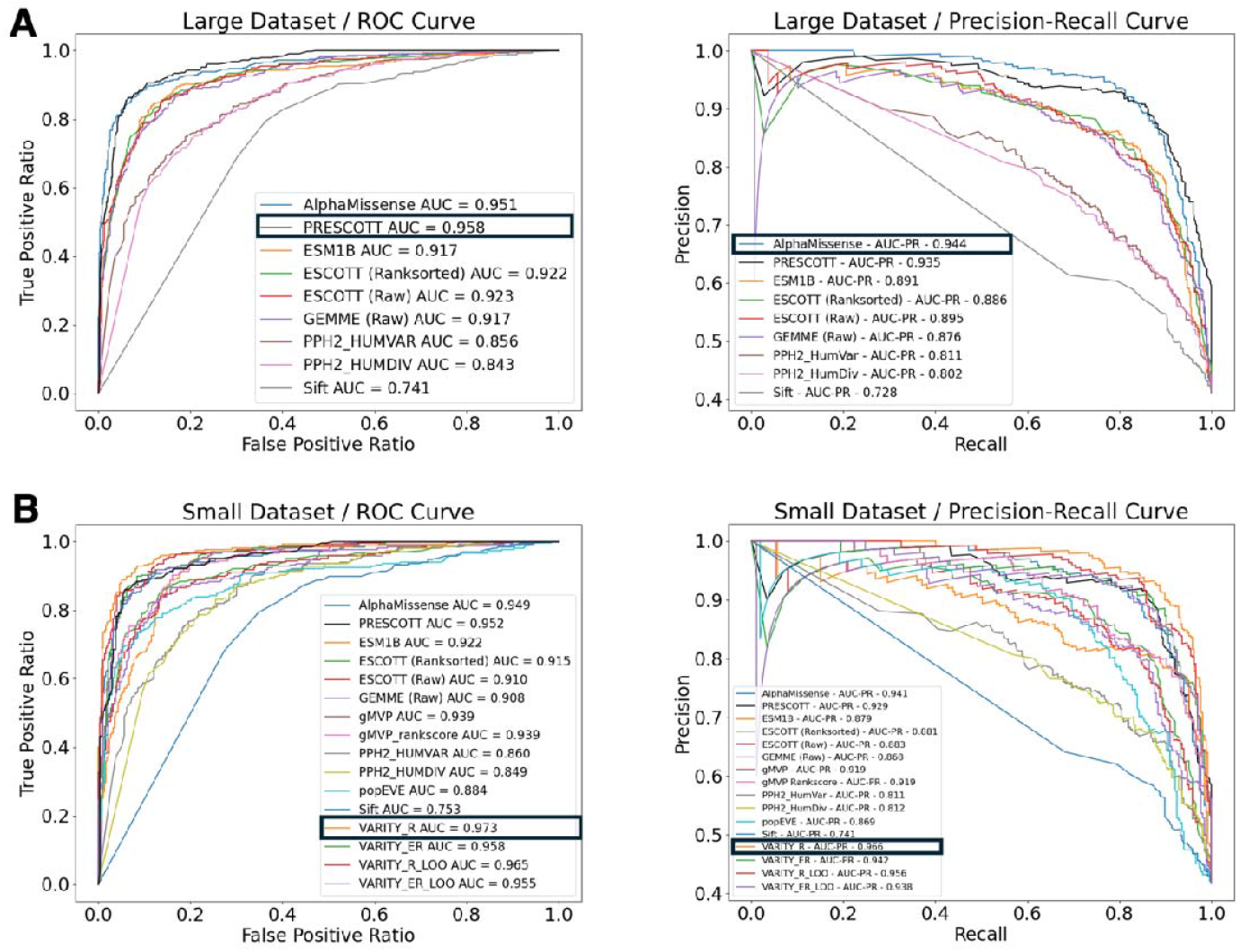
ROC and Precision-Recall Curves for the large and small datasets. A) Left panel: ROC curves and AUC values of each method for the large dataset. Right panel: Precision-Recall curves and AUC-PR values for the large dataset. B) Left panel: ROC curves and AUC values of each method for the small dataset. Right panel: Precision-Recall curves and AUC-PR values for the small dataset.

#### APE Score Performance for Binary Labeled Mutants

APE score is a linear combination score of AlphaMissense, PRESCOTT and ESM1b. To measure APE score performance for distinguishing pathogenic and benign variants, we plotted ROC and Precision-Recall curves for the large and small datasets in **Fig. *4***. AUC and AUC-PR values are 0.968 and 0.962 for the large dataset, respectively (**Fig. *4*** A). These values are greater than the prediction performances of each individual predictor. When the small dataset is investigated, we obtain AUC as 0.966 and AUC-PR as 0.961 (**Fig. *4*** B). Similar to the large dataset, we observe that the combined APE score has a higher accuracy than its components.

**Fig. 4.**
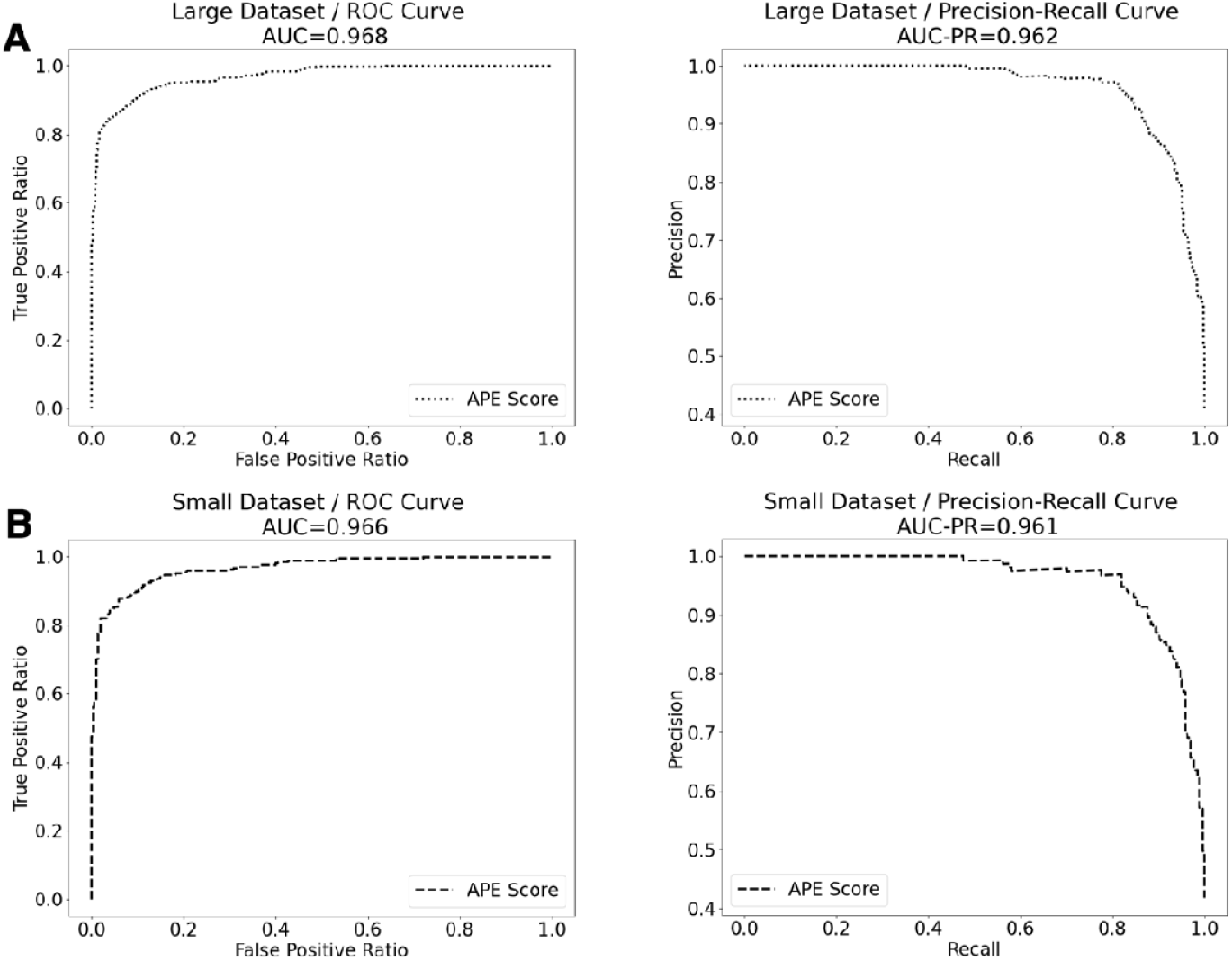
ROC and Precision-Recall curves of APE score for the large and the small datasets. A) Left panel: ROC curve (dotted line) and AUC values of APE score for the large dataset. Right panel: Precision-Recall curve (dotted line) and AUC-PR values of APE score for the large dataset. B) Left panel: ROC curve (dashed line) and AUC values of APE score for the small dataset. Right panel: Precision-Recall curve (dashed line) and AUC-PR values of APE score for the small dataset.

A key question may arise about why we used a linear combination of the three state-of-the-art methods instead of their pairwise linear combinations. Due to this reason, we investigated all pairwise combinations of AlphaMissense, PRESCOTT and ESM1b, and calculated AUC/AUC-PR scores. Our results demonstrates that triple combination is better than all possible pairwise combinations in terms of AUC and AUC-PR values (see Fig. S4). Furthermore, there is not a consistency for the performance of the pairs. While AlphaMissense+PRESCOTT becomes the best pair in the large dataset, ESM1b+PRESCOTT becomes the best in the small dataset. As a result, APE score should be chosen over pairwise combinations of three methods.

#### A New Variant Classification based on APE Scores

In previous subsections, we demonstrated that APE scores give better missense mutational effect predictions in terms of AUC and AUC-PR. However, we still don’t know upper and lower limits of APE scores for pathogenic and benign variant classification. Therefore, it is essential to plot distributions of APE scores for pathogenic and benign labeled variants. Both for the large and the small datasets, majority of pathogenic variants have APE scores greater than 0.6. On the other hand, majority of benign variants have an APE score in the range of [0.0, 0.4] (**Fig. *5*** A and B). Even though this crude analysis gives some information on how to classify variants according to their APE scores, we wanted to carry out a more systematic classification of the variants into three classes: pathogenic, variants of unknown significance (VUS) and benign. We followed a protocol that has been used in a previous study (Cheng et al. 2023). We plotted precision and recall curves for various APE score thresholds. Then, we found intersection points of precision and recall curves with 90% probability line. The intersection value of 90% probability line with recall curve determines upper boundary of benign class, while intersection value of 90% probability line with precision curve determines lower boundary of pathogenic class. Accordingly, upper limit of benign variants is 0.42 for the large dataset, while it is 0.44 for the small dataset. Similarly, lower limit of APE scores for pathogenic classification is 0.48 for the large dataset and 0.50 for the small dataset (**Fig. *5*** C and D). All the values remaining between these ranges should be classified as VUS. Despite little differences in the boundaries for the large and small datasets, the results remain consistent with our observations in (**Fig. *5*** A and B)

**Fig. 5.**
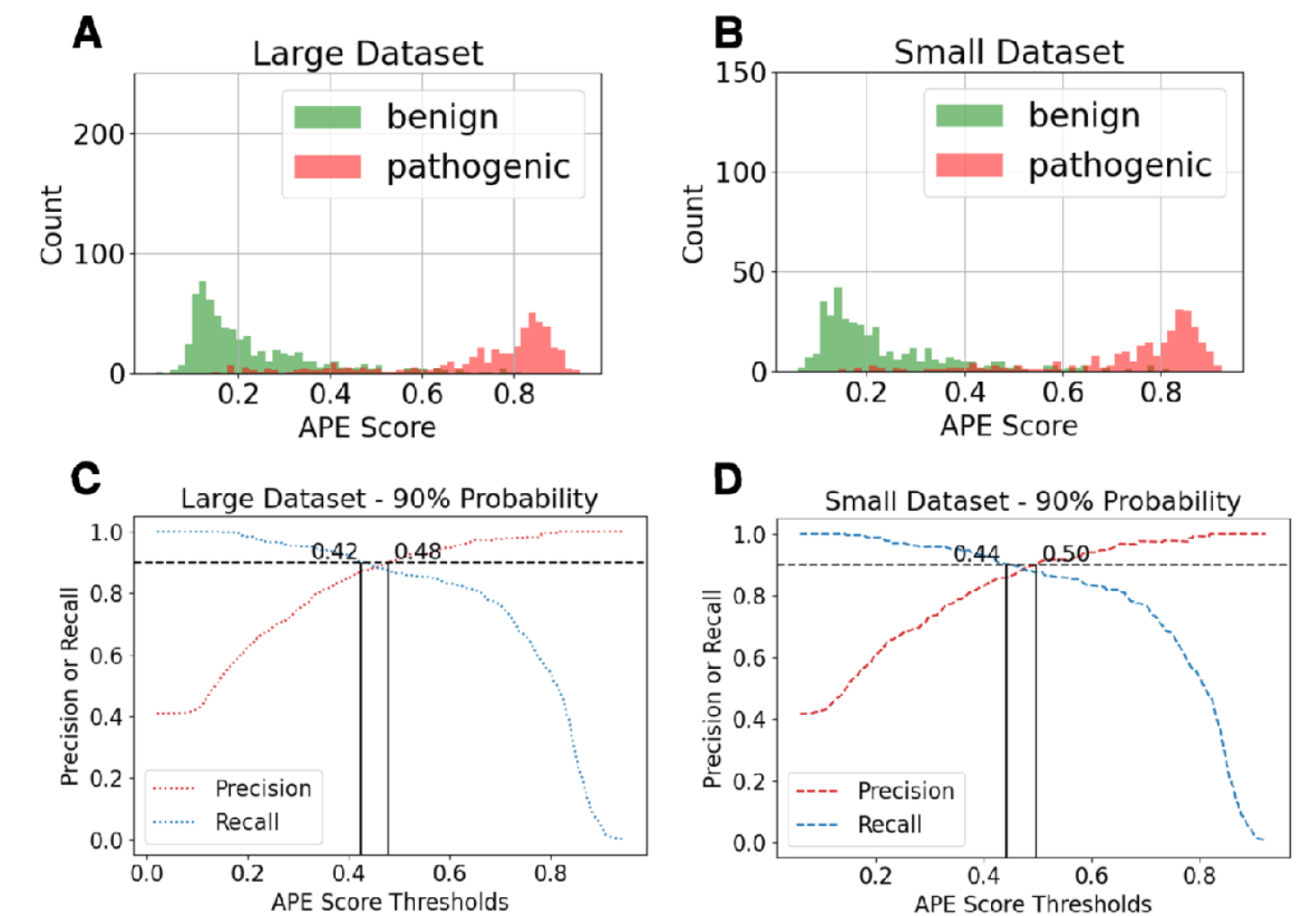
Score distributions and Precision/Recall Curves with respect to different APE score values. A) APE score distributions of benign and pathogenic mutations in the large dataset. B) APE score distributions of benign and pathogenic mutations in the small dataset. C) Precision and recall curves for different APE score thresholds in the large dataset (dotted colored curves). Horizontal black dashed line is 90% probability line. The recall (blue dotted) curve intersects with 90% probability line at 0.42, while the precision (red dotted) curve intersects with the 90% probability line at 0.48 in the large dataset. D) Precision and recall curves for different APE score thresholds in the small dataset (dashed colored curves). Horizontal black dashed line is 90% probability line. The recall (blue dashed) curve intersects with 90% probability line at 0.44, while the precision (red dashed) curve intersects with the 90% probability line at 0.50 in the small dataset.

#### Measuring Performance of APE Classification

We wanted to see performance of APE classification on the large and small datasets. Therefore, we classified pathogenic and benign labeled mutations according to the boundaries determined in the previous subsection. Out of 438 pathogenic mutations in the large dataset, only 15 variants were classified as VUS and only 42 variants were classified as benign (**Fig. *6*** A). On the other hand, only 42 mutations were classified as pathogenic and 18 variants were classified as VUS out of 630 benign labeled variants in the large dataset (**Fig. *6*** B). When these APE score classifications are compared with AlphaMissense, PRESCOTT and ESM1b, we observe that main benefit of APE classification lies in its reduction of misclassified benign mutations. The results for the small dataset, given in **Fig. *6*** C and D support this claim.

**Fig. 6.**
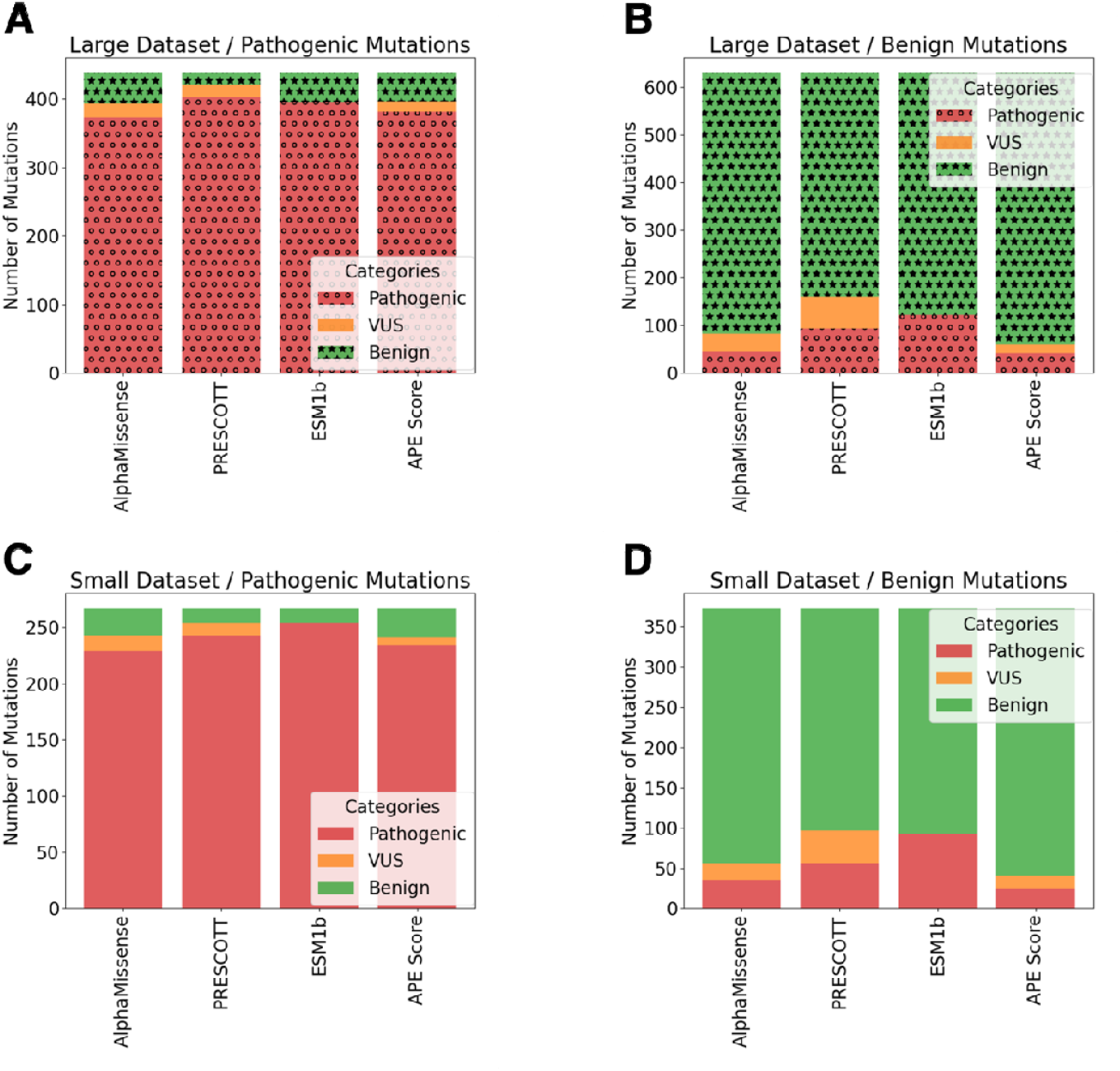
Comparison of predicted classifications of AlphaMissense, PRESCOTT, ESM1b and APE score with experimental/clinical classifications. Green color shows benign mutations, orange color shows variants of unknown significance (VUS) and red color shows pathogenic mutations. A) Classifications of AlphaMissense, PRESCOTT, ESM1b and APE score methods for pathogenic mutations in the large dataset. B) Classifications of AlphaMissense, PRESCOTT, ESM1b and APE score methods for benign mutations in the large dataset. C) Classifications of AlphaMissense, PRESCOTT, ESM1b and APE score methods for pathogenic mutations in the small dataset. D) Classifications of AlphaMissense, PRESCOTT, ESM1b and APE score methods for benign mutations in the large dataset.

#### APE Scores and Classifications for All Variants

In this work, we demonstrated that a combined score, named APE score, can predict missense mutational effect better than its three state-of-the art components over 9 deep mutational scanning experiments and a set of 1068 variants in 24 oncoproteins. However, there are thousands of variants without any score or classification in these oncoproteins. Due to this reason, we calculated APE scores and APE classifications for all variants in these proteins as a service to the experimental and clinical communities. We used [0.0-0.42] as benign classification range for APE scores. Since minimal value for pathogenic APE classification was 0.48 in the large dataset and 0.50 in the small dataset, we decided to use 0.50 as the lower limit of pathogenic classification (**Fig. *7*** A). This approach enlarges VUS range but tries to ensure a safer classification for the likely pathogenic variants. In total, there are 444087 variants in these 24 proteins. As a result of APE classification, 180994 variants were classified as ‘Likely pathogenic’, 220726 variants were classified as ‘Likely benign’ and finally 42367 variants were classified as VUS (**Fig. *7*** B). The data about each variant in all proteins can be found at https://doi.org/10.5281/zenodo.15006566.

**Fig. 7.**
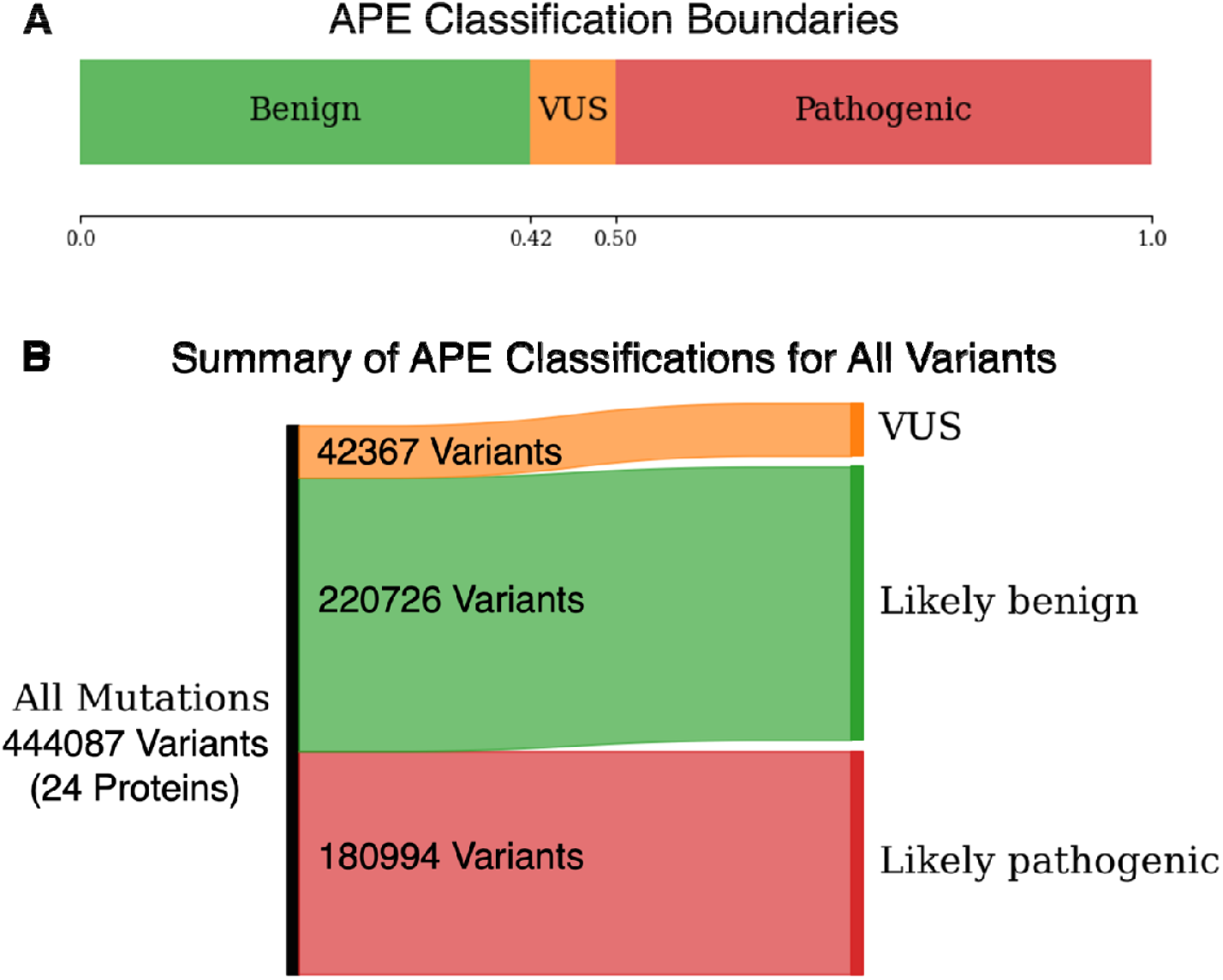
A) APE classification boundaries used to classify all variants from 24 proteins. B) A summary of classification results for 444087 variants from 24 proteins in the large dataset.

## Conclusions

In this study, we proposed a new collaborative metric called APE score, which is a linear combination of three unsupervised state-of-the-art variant effect prediction algorithms. We demonstrated its accuracy on nine deep mutational scanning experiments from five oncoproteins. We assessed its binary classification performance on a large independent dataset of 1068 pathogenic and benign variants. We removed some proteins from the large dataset to demonstrate validity of our claims in a smaller dataset. Furthermore, we showed that APE score is better than pairwise combinations of its three constituents. Our analysis reveals that APE score reduces number of VUS and pathogenic labels in benign class, and therefore, provides a higher accuracy in terms of three metrics, namely Spearman correlation, AUC and AUC-PR.

Of course, using a combination of different methods is not a novel idea. There have been numerous studies incorporating results of various methods, mostly in supervised learning algorithms. For example, REVEL is an ensemble method utilizing results of thirteen other methods as features in a supervised learning algorithm (Ioannidis et al. 2016). ClinPred is another supervised ensemble learning method that incorporates results from various methods (Alirezaie et al. 2018). CADD combines 63 functional annotations, including annotations from supervised methods like PolyPhen2, into a single score (Kircher et al. 2014; Rentzsch et al. 2019). In our case, we selected AlphaMissense, PRESCOTT and ESM1b because they are not supervised learning algorithms. Since cancer proteins are widely investigated, they are commonly used in almost all supervised learning algorithms for training, which causes a bias in predicting variant effects in oncoproteins. As a result, our three methods are not prone to this bias problem. In conclusion, APE score components are not supervised methods that may have biases due to the data in the training set and these components have never been used together before.

## Data Availability

The data used and generated in this work is deposited at https://doi.org/10.5281/zenodo.15006566.

## Data Availability

The data used and generated in this work is deposited at https://doi.org/10.5281/zenodo.15006566.

## Competing interests

The authors declare that they have no conflict of interest.

## Author Contributions

The authors contributed equally to this work.

## Notes

### Competing Interest Statement

The authors have declared no competing interest.

### Funding Statement

This study did not receive any funding.

