## Supplementary Information for "A Collaborative Approach to Improving Missense Mutational Effect Predictions in Oncoproteins"

Keywords

**This Supplementary Information contains 4 figures and 1 table.**

FIGURES

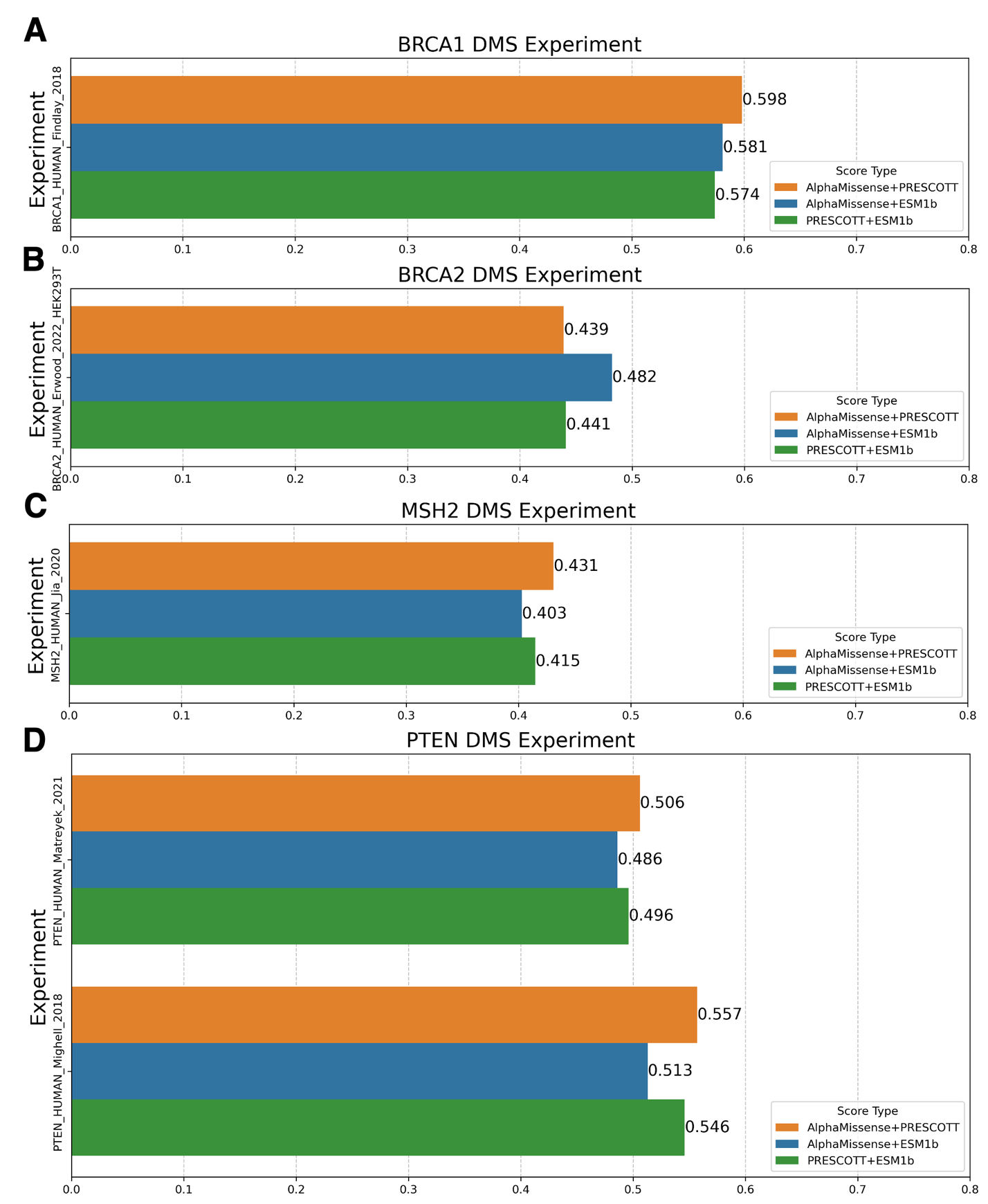

Figure S 1. Comparison of Spearman correlation performances for pairwise combinations of AlphaMissense, PRESCOTT and ESM1b across various deep mutational scanning experiments A) BRCA1 B) BRCA2 C) MSH2 D) PTEN.

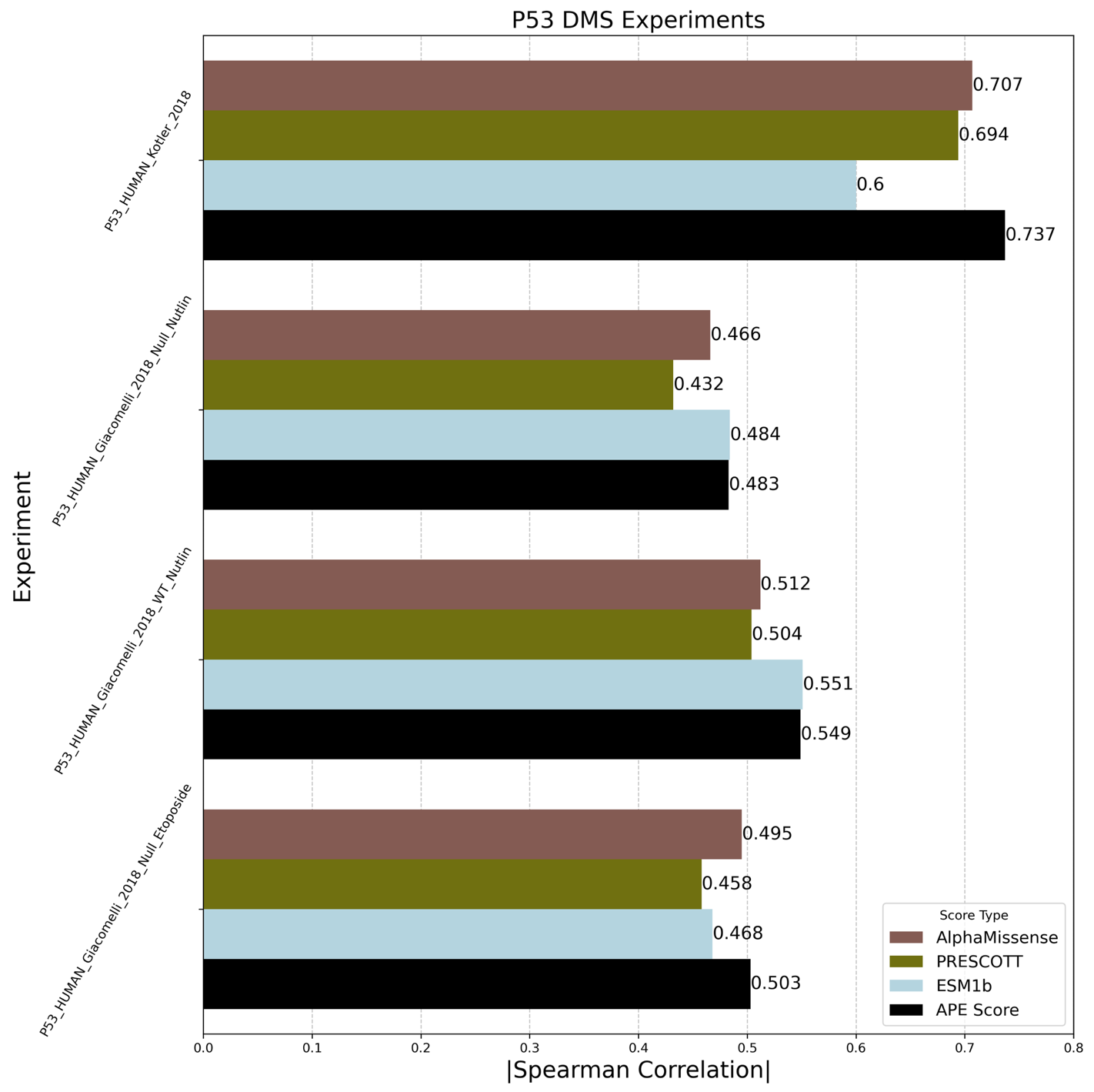

Figure S 2. Comparison of AlphaMissense (brown bars), PRESCOTT (olive bars) and ESM1b (light blue bars) Spearman correlation performances with APE score performance across various deep mutational scanning experiments of P53.

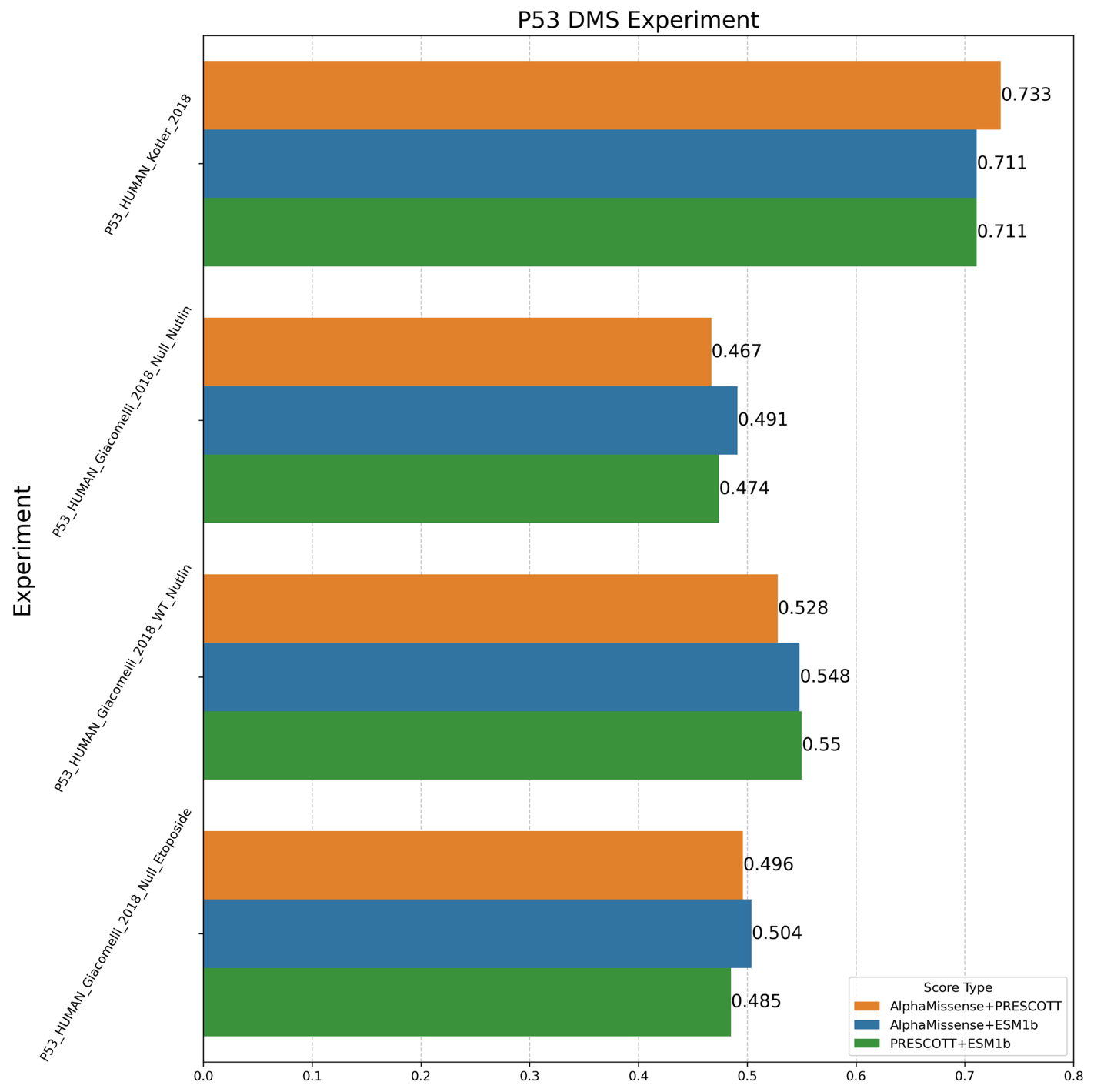

Figure S 3. Comparison of Spearman correlation performances of pairwise combinations of AlphaMissense, PRESCOTT and ESM1b across various deep mutational scanning experiments of P53.

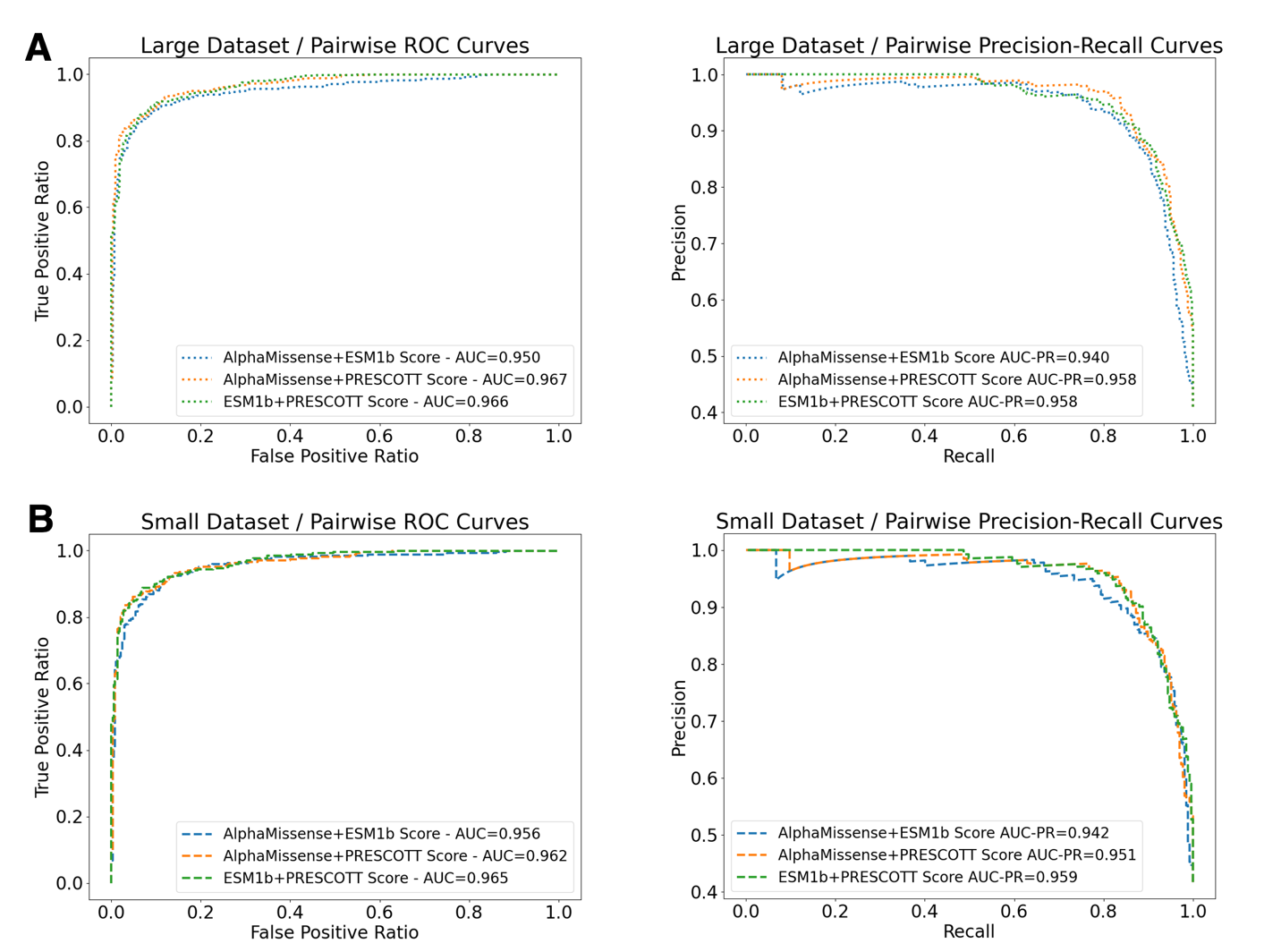

Figure S 4. ROC and Precision-Recall Curves of the pairwise combinations of three methods for the large and small datasets. A) Left panel: ROC curve (dotted line) and AUC values of the pairwise combinations of three methods for the large dataset. Right panel: Precision-Recall curve (dotted line) and AUC-PR values of the pairwise combinations of three methods for the large dataset. B) Left panel: ROC curve (dashed line) and AUC values of the pairwise combinations of three methods for the small dataset. Right panel: Precision-Recall curve (dashed line) and AUC-PR values of the pairwise combinations of three methods for the small dataset.

TABLES

Table S1. Number of amino acids in each protein and number of variants in each deep mutational scanning experiment.

| **Experiment** | **Number of Amino Acids** | **Number of Variants in the Experiment** |
| --- | --- | --- |
| **BRCA1_HUMAN_Findlay_2018** | 1863 | 1837 |
| **BRCA2_HUMAN_Erwood_2022_HEK293T** | 3418 | 265 |
| **MSH2_HUMAN_Jia_2020** | 934 | 16749 |
| **PTEN_HUMAN_Matreyek_2021** | 403 | 5083 |
| **PTEN_HUMAN_Mighell_2018** | 403 | 7260 |
| **P53_HUMAN_Kotler_2018** | 393 | 1048 |
| **P53_HUMAN_Giacomelli_2018_Null_Etoposide** | 393 | 7467 |
| **P53_HUMAN_Giacomelli_2018_Null_Nutlin** | 393 | 7467 |
| **P53_HUMAN_Giacomelli_2018_WT_Nutlin** | 393 | 7467 |
